# Rationale and design of the Learning Implementation of Guideline-based decision support system for Hypertension Treatment (LIGHT) Trial and LIGHT-ACD Trial

**DOI:** 10.1101/2021.03.11.21253427

**Authors:** Jiali Song, Xiu-Ling Wang, Bin Wang, Yan Gao, Jia-Min Liu, Hai-Bo Zhang, Xi Li, Jing Li, Ji-Guang Wang, Jun Cai, Jeph Herrin, Jane Armitage, Harlan M. Krumholz, Xin Zheng, on behalf of the LIGHT Collaborative Group

## Abstract

**Background:** Computerized clinical decision support systems (CDSS) are low-cost, scalable tools with the potential to improve guideline-based antihypertensive treatment in primary care, but their effectiveness needs testing in pragmatic trials.

**Methods:** The Learning Implementation of Guideline-based decision support system for Hypertension Treatment (LIGHT) trial is a pragmatic, four-stage, cluster-randomized trial conducted in 94 primary care practices in China. For each city-based stage, practices are randomly assigned to either implementation of the CDSS for hypertension management (which guides physicians treatment recommendations based on measured blood pressure and patient characteristics), or usual care. Patients are enrolled during the first 3 months after site randomization and followed for 9 months. The primary outcome is the proportion of hypertension management visits at which guideline-based treatment is provided.

In a separate sub-study conducted within the CDSS, with the patient as the unit of randomization, the LIGHT-ACD trial, patients are randomized to receive different initial mono- or dual- antihypertensive therapy. The primary outcome of the LIGHT-ACD trial is the change in blood pressure from the first visit after site randomization to 9 months.

**Discussion:** The LIGHT trial will provide evidence on the effectiveness of a CDSS for improving guideline adherence for hypertension management in primary care in China. The sub-study, LIGHT-ACD trial, will provide data on the effect of different initial antihypertensive regimens for blood pressure management in this setting.

**Trial registration:** ClinicalTrials.gov, identifier: LIGHT (NCT03636334) and LIGHT-ACD (NCT03587103). Registered on 3 July 2018.

## BACKGROUND

Hypertension is the leading modifiable risk factor for death globally.^1^ Over the past decades, the number of individuals with hypertension is estimated to have increased by 90%, with the majority of the increase occurring in low- and middle-income countries.^2^ In China, an estimated 244.5 million adults have hypertension, and only about 15% of these individuals have adequate blood pressure control, resulting in major health and economic burdens.^3^

Improving the performance of primary care providers, who play a key role in managing hypertension, and ensuring their adherence to guideline-recommended antihypertensive treatments are public health priorities in China.^4^ Despite decade-long efforts to improve the primary care system,^5^ there are quality issues^3, 6^, and decision support tools may help bridge this gap.^4, 7^ Traditional strategies to improve care, including training sessions have only yielded modest effects.^8^ Furthermore, such interventions are often difficult to implement widely because retraining of providers is resource-intensive.^4, 7, 9^ The barriers to adequate management of hypertension in China have led to calls for the implementation of computerized clinical decision support system (CDSS) to improve performance and aid compliance with guidelines.^4, 9^ Such systems are characterized by computerized algorithms that generate guideline-based recommendations, and hence have the potential to improve appropriate medication prescribing. However, prior studies assessing the effectiveness of CDSS have shown mixed results.^10-13^ A scalable CDSS integrated into primary care settings could improve the performance of providers and mitigate the burden of hypertension in China and other countries.

In addition, in the course of testing the CDSS, there is an opportunity to test different regimens. Blood pressure guidelines endorse a range of therapies as equivalent even though there are a paucity of head-to-head comparisons. Treatment options supported by previously published clinical trials or meta-analysis include angiotensin-converting enzyme (ACE) inhibitors/angiotensin receptor blockers (ARB), calcium channel blockers (CCB), or thiazide/thiazide-like diuretics.^14, 15^ Although the blood pressure lowering effects of these antihypertensive medication classes are well documented, there are differences in individual tolerability and drug-drug interactions.^14, 16, 17^ The CDSS pragmatic trial to promote guideline adherence can rotate medication recommendations so that there is a nested trial comparing different guideline-endorsed regimens.

Accordingly, we developed a CDSS for hypertension management and designed a pragmatic, cluster-randomized controlled trial, the Learning Implementation of Guideline-based decision support system for Hypertension Treatment (LIGHT) trial, to assess its effectiveness for improving antihypertensive management in primary care in China. Within the framework of LIGHT trial, the LIGHT-ACD trial aims to test alternative, guideline-recommended regimens for blood pressure reduction.

## METHODS/DESIGN

### The LIGHT trial

The LIGHT trial is a pragmatic, parallel-group, four-stage, cluster-randomized controlled trial assessing the effectiveness of CDSS, with primary care practices as the unit of randomization. (**Supplement 1**) The protocol is reported according to the Standard Protocol Items: Recommendations for Interventional Trials (SPIRIT) checklist^18^ (**Additional file 1**).

In each stage, up to 36 urban primary care practices were to be randomized to having the CDSS embedded into their existing electronic health care record (EHR) or to continue use of the EHR without the CDSS. During the first three months after randomization, patients who attend the clinic for hypertension management are screened for eligibility (see below) and if eligible asked to attend the clinic at least every 3 months with data collected at each visit.^19^ (**Figure 2**)

Urban primary care practices with hypertension clinics were eligible to be included if: (1) at least one agent from each of the four classes of antihypertensive medication was available for prescription (ACE-inhibitor/ARB, beta-blocker, CCB, thiazide/thiazide-like diuretic); (2) an EHR was being used for hypertension management providing a structured template for collection of patient data; and (3) at least 100 patients with hypertension were routinely being seen. Primary care practices selected for each stage were randomized to the intervention (CDSS) or control arm (usual care) in a 1:1 stratified by the baseline appropriate (guideline-based) antihypertensive treatment rates and site characteristics (**Supplement 2**).

#### The Computer Decision Support System (CDSS)

The CDSS is a software algorithm which was developed by a multidisciplinary team including clinicians and information technology experts. The CDSS uses information recorded in the EHR and measurements taken at the visit to make recommendations about the antihypertensive treatment for the patient. The recommendations are based on the Chinese primary care hypertension guidelines^19^ but are generally consistent with international guidance^20, 21^ and include a recommendation to add one or more of the 4 classes of medication at half or full dose (**Supplement 4**). The logic of the CDSS was tested using simulated patient data to trigger each possibility. In order to ensure that the intervention is being reliably delivered to and used by doctors, clicking the icon is designed to be mandatory before documentation of the prescription. If the doctor decides not to follow the recommendation, a pop-up message stating “your prescription did not match with CDSS recommendation, please re-evaluate the classes/doses/frequencies of drugs.” and prescriptions can be modified accordingly. If the doctor decides not to follow the CDSS recommendation, relevant reasons (e.g., doctor preference or patient refusal) is recorded. In addition, if prescriptions involve contraindicated drugs, under-dosing, or over-dosing, a message is also triggered.

In control sites the EHR is used to record the same information but no treatment recommendation is made and prescribing decisions are made by doctors based on their knowledge and usual practice and recorded in the EHR.

#### Eligibility

Local residents aged ≥18 years with established hypertension attending the clinic are eligible for the LIGHT trial if they are taking 0–2 classes of antihypertensive medication. Major exclusion criteria include: (1) history of coronary heart disease, heart failure, or chronic kidney disease; or (2) intolerance to ≥2 classes of antihypertensive medications (**Table 1**). As the visit is the observation unit for the primary and secondary outcomes, visits involving treatment of hypertension (hypertension visits) of eligible patients are included in the analysis (**Supplement 3**).

**Table 1.**
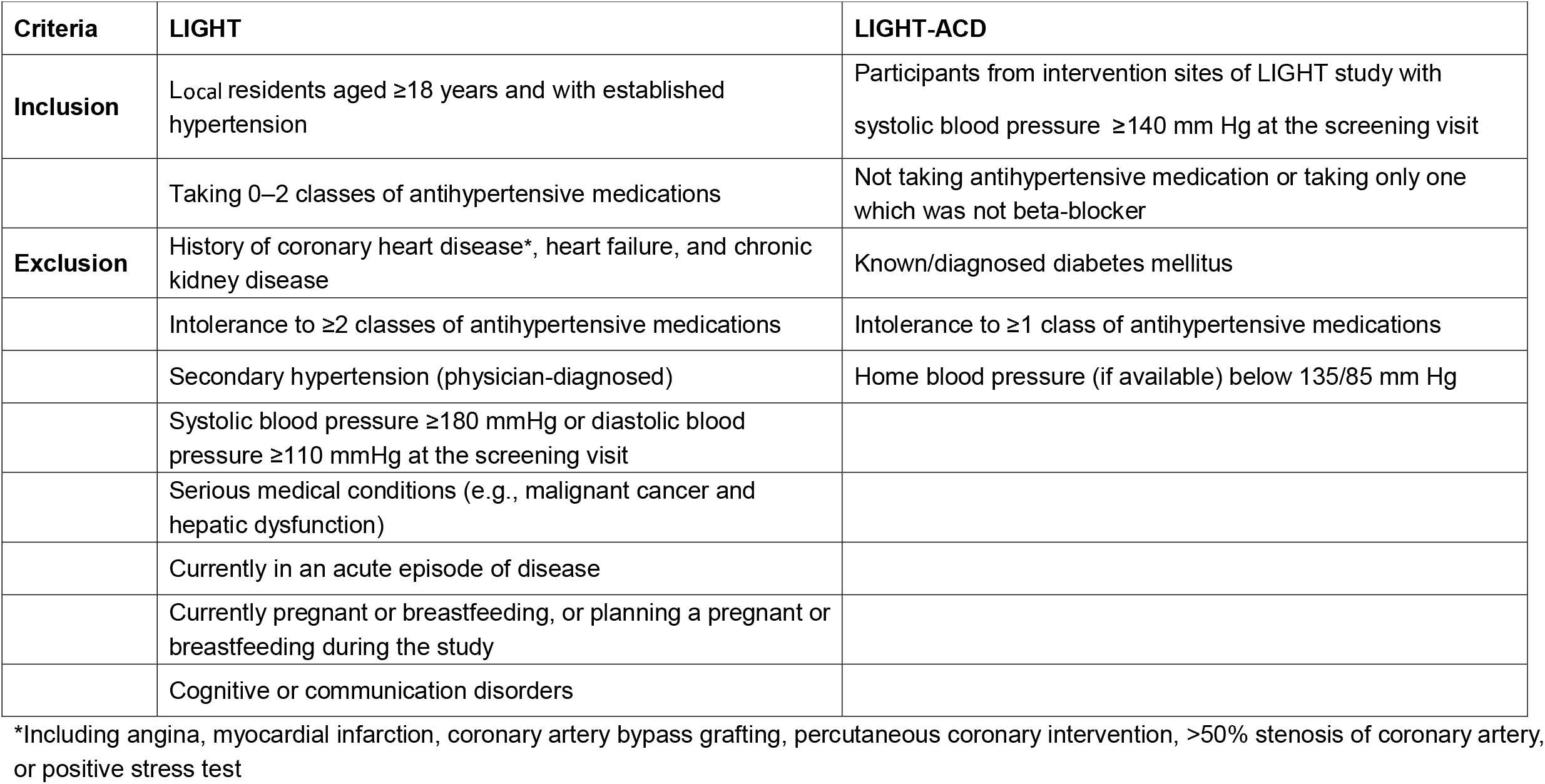
Inclusion and exclusion criteria of participants for LIGHT and LIGHT-ACD trials.

| **Criteria** | **LIGHT** | **LIGHT-ACD** |
| --- | --- | --- |
| **Inclusion** | Local residents aged ≥18 years and with established hypertension | Participants from intervention sites of LIGHT study with systolic blood pressure ≥140 mm Hg at the screening visit |
|  | Taking 0–2 classes of antihypertensive medications | Not taking antihypertensive medication or taking only one which was not beta-blocker |
| **Exclusion** | History of coronary heart disease*, heart failure, and chronic kidney disease | Known/diagnosed diabetes mellitus |
|  | Intolerance to ≥2 classes of antihypertensive medications | Intolerance to ≥1 class of antihypertensive medications |
|  | Secondary hypertension (physician-diagnosed) | Home blood pressure (if available) below 135/85 mm Hg |
|  | Systolic blood pressure ≥180 mmHg or diastolic blood pressure ≥110 mmHg at the screening visit |  |
|  | Serious medical conditions (e.g., malignant cancer and hepatic dysfunction) |  |
|  | Currently in an acute episode of disease |  |
|  | Currently pregnant or breastfeeding, or planning a pregnant or breastfeeding during the study |  |
|  | Cognitive or communication disorders |  |

\*Including angina, myocardial infarction, coronary artery bypass grafting, percutaneous coronary intervention, >50% stenosis of coronary artery, or positive stress test

#### Data collection, quality control, and management

To improve the workflow patient data are collected via the customized EHR, which only collects information necessary for hypertension management (**Table 2**). Blood pressure is measured in the sitting position after at least a 5-minute rest, with a validated automated sphygmomanometer (Omron HBP-1300)^22^. Two blood pressure readings are taken 1–2 minutes apart and the average value is recorded.

**Table 2.** Data elements collected in baseline, recruitment, and follow-up.

|  | **Baseline** | **Recruitment** | **Follow-up** |
| --- | --- | --- | --- |
| Socio-demographics<br>(age, gender, education, and insurance) | √ | √ | √ |
| Physical measurements<br>(blood pressure, heart rate, waist, height, and weight) | √ | √ | √ |
| Self-reported home monitoring blood pressure | √ | √ | √ |
| Cardiovascular risk factors | √ | √ | √ |
| Co-morbidities | √ | √ | √ |
| Hospitalizations |  | √ | √ |
| Adverse events |  | √ | √ |
| Current medications | √ | √ | √ |
| Medication adherence* | √ | √ | √ |
| Side-effects related to antihypertensive medication |  | √ | √ |
| CDSS recommendations<sup>#</sup> |  | √ | √ |
| Prescriptions | √ | √ | √ |
| Reasons for not following CDSS<sup>#</sup> |  | √ | √ |

\*Adherence to each antihypertensive medication is collected as three classes: regularly, intermittently, and rarely
<sup>#</sup>Only in the intervention sites of LIGHT trial

A bespoke website was developed to monitor progress and quality of the trial in real time. The CDSS recommendations are regularly reviewed via the website to ensure that the CDSS is working as designed. In addition, on-site audit of data collection is regularly conducted by the research team. To ensure the accuracy of documentation of the blood pressure values, values in the EHR are randomly selected (at least one value per site) and checked on a daily basis against the recordings in the blood pressure monitor.

All data are securely transmitted to the central server through automatic electronic transfer and securely stored in an encrypted and password-protected database. Data confidentiality policies on data collection, storage, and analysis have been strictly imposed in order to ensure the confidentiality of personal information.

#### Outcomes

The primary outcome for the LIGHT trial is the proportion of hypertension visits during which appropriate antihypertensive treatment is prescribed. Appropriate antihypertensive treatment is defined as a prescription compliant with the pre-specified guideline-based recommendations (see detailed recommendations specifications in **Supplement 5**).

The secondary outcomes include the average change in systolic blood pressure, the proportion of those whose blood pressure is controlled at 9 months, and the proportion of hypertension visits with *acceptable* antihypertensive treatment, which is defined as either appropriate antihypertensive treatment (as above) or those treatments with acceptable reasons for failing to titrate antihypertensive treatment. An exploratory outcome is the number of patients experiencing a vascular event defined as a composite of cardiac death, non-fatal stroke, and non-fatal myocardial infarction. (**Table 3**)

**Table 3.**
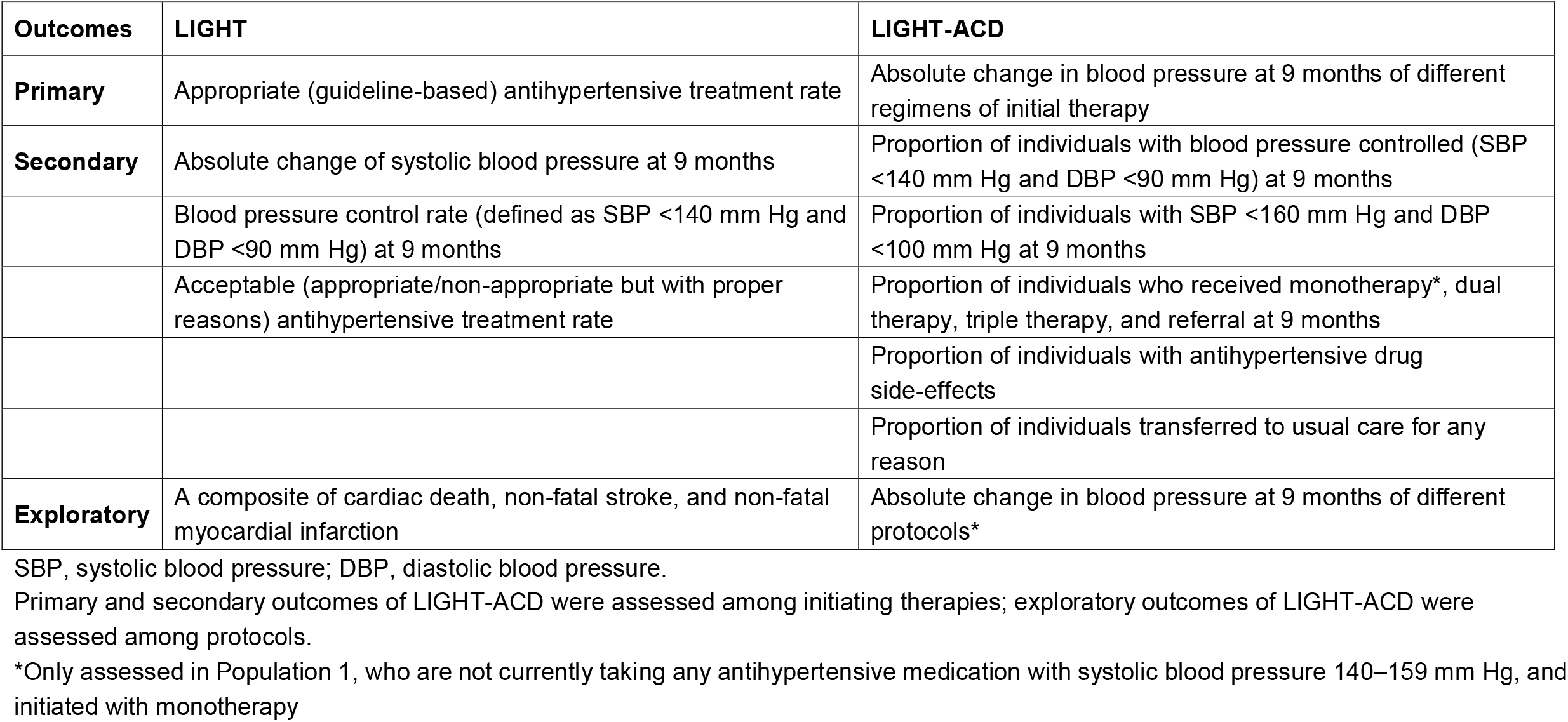
Outcomes of LIGHT and LIGHT-ACD trials.

| **Outcomes** | **LIGHT** | **LIGHT-ACD** |
| --- | --- | --- |
| **Primary** | Appropriate (guideline-based) antihypertensive treatment rate | Absolute change in blood pressure at 9 months of different regimens of initial therapy |
| **Secondary** | Absolute change of systolic blood pressure at 9 months | Proportion of individuals with blood pressure controlled (SBP <140 mm Hg and DBP <90 mm Hg) at 9 months |
|  | Blood pressure control rate (defined as SBP <140 mm Hg and DBP <90 mm Hg) at 9 months | Proportion of individuals with SBP <160 mm Hg and DBP <100 mm Hg at 9 months |
|  | Acceptable (appropriate/non-appropriate but with proper reasons) antihypertensive treatment rate | Proportion of individuals who received monotherapy*, dual therapy, triple therapy, and referral at 9 months |
|  |  | Proportion of individuals with antihypertensive drug side-effects |
|  |  | Proportion of individuals transferred to usual care for any reason |
| **Exploratory** | A composite of cardiac death, non-fatal stroke, and non-fatal myocardial infarction | Absolute change in blood pressure at 9 months of different protocols* |

SBP, systolic blood pressure; DBP, diastolic blood pressure.
Primary and secondary outcomes of LIGHT-ACD were assessed among initiating therapies; exploratory outcomes of LIGHT-ACD were assessed among protocols.
\*Only assessed in Population 1, who are not currently taking any antihypertensive medication with systolic blood pressure 140–159 mm Hg, and initiated with monotherapy

#### Sample size

As this is a staged pragmatic trial, we have the capacity to add sites as the trial progresses. Initial statistical power was based on the number of sites potentially eligible in stage 1 at the time the power analysis was being conducted; thus, we assumed that at least 10 primary care practices would be randomized to the intervention arm and 10 to the control arm, the baseline appropriate treatment rate would be 55% with maximum type II error of α=0.05, a moderate intra-site correlation of 0.05, a within-patient correlation of 0.1, and statistical power of 90%, we needed 3 hypertension visits per patient for 50 patients at each site in order to detect a 18% absolute difference in appropriate treatment rate between the two arms. Subsequent enrollment increased the number of participating sites to 94, randomized in 4 waves; under the same assumptions as above we anticipate that the final sample will provide 90% power to detect a true difference of 4% in appropriate treatment rate. This difference is close to the average effect of CDSS in improving process of care in prior studies.^23^

#### Statistical analysis

The analyses and reporting of the results will follow the Consolidated Standards of Reporting Trials guidelines for cluster randomized controlled trials.^24^ All the analyses will be performed on an intention-to-treat basis. Multiple imputation by chained equations will be used to account for missing values.

With all comparative outcomes, absolute differences between the intervention and control sites with 95% CIs will be presented. Implementation stages will be treated as strata, with adjustment for calendar time to account for secular trends. The analysis of both primary and secondary outcomes will account for the clustering effect using mixed-effects models with site as a random effect. The consistency of treatment effects on the primary outcome will be explored in predefined subgroups, including age, gender, education, implementation stage, and tertile of cluster-level endpoints. All statistical tests will be performed using 2-sided tests at the 0.05 level of significance but the number of tests and p value will be taken into account in the interpretation of the results

### The LIGHT-ACD trial

The LIGHT-ACD trial is an individually randomized sub-study embedded into the intervention sites of LIGHT trial. In this sub-study, patients are randomized to receive various initial antihypertensive therapies and blood pressure changes between different regimens are compared (**Figure 2**).

#### Recruitment

The patients at the intervention sites of LIGHT trial are eligible if they have a measured baseline SBP ≥ 140 mm Hg, and are taking 0–1 class of antihypertensive medication. Key exclusion criteria include diabetes mellitus and intolerance to at least one of the four class of antihypertensive medications (**Table 1**). The eligible patients in the LIGHT-ACD trial are categorized into 2 subpopulations. Patients with a SBP of 140–159 mm Hg who are not taking any antihypertensive medication are categorized as Population 1, the remainder as Population 2.

#### Randomization

Populations 1 and 2 are randomized separately (**Figure 1**). Population 1 individuals are randomized to receive one of the initial monotherapies of A (ACE-inhibitor or ARB), C (CCB), or D (diuretics). If additional treatment is needed, they are further randomized to add either C or D so patients initiated with A are follow either A-AC-ACD or A-AD-ADC with C and D as the add-on medications, respectively. The randomization of protocols among patients initiated with C or D is similar to patients initiated with A. Population 2 are randomized to receive one of the three initial dual therapies of AC, AD, or CD. Subsequently, D, C, or A is added to achieve blood pressure control, if necessary, respectively (**Figure 1**). Minimization randomization is used to ensure balance on age, gender and education level among the three arms of each populations.^25^ Neither patients nor physicians are blinded to treatment allocation but the allocation is concealed within the CDSS.

**Figure 1.**
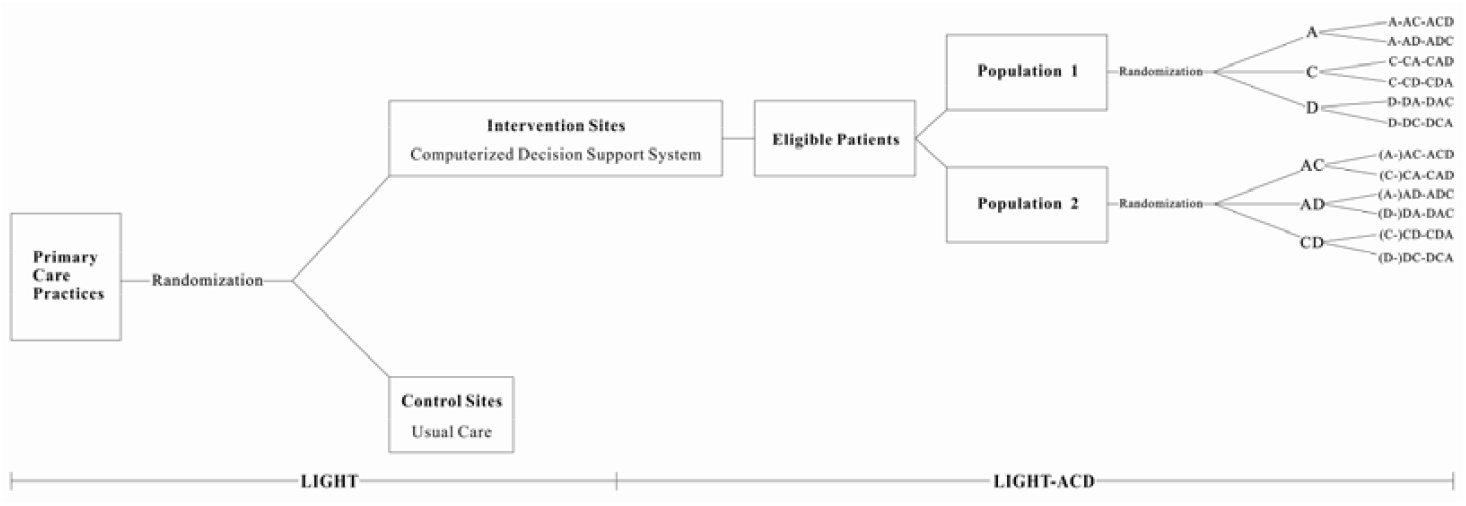
Infrastructure of the LIGHT and LIGHT-ACD trials. Population 1: Participants with systolic blood pressure (SBP) of 140–159 mm Hg, and were not taking any antihypertensive medication. Population 2: Participants with SBP ≥160 mm Hg and were not taking any antihypertensive medication or taking one antihypertensive medication which was not beta-blocker, or those with SBP 140–159 mm Hg and were taking one antihypertensive medication which was not beta-blocker. A: angiotensin-converting enzyme inhibitor or angiotensin receptor blocker; C: calcium channel blocker; D: diuretic.

**Figure 2.**
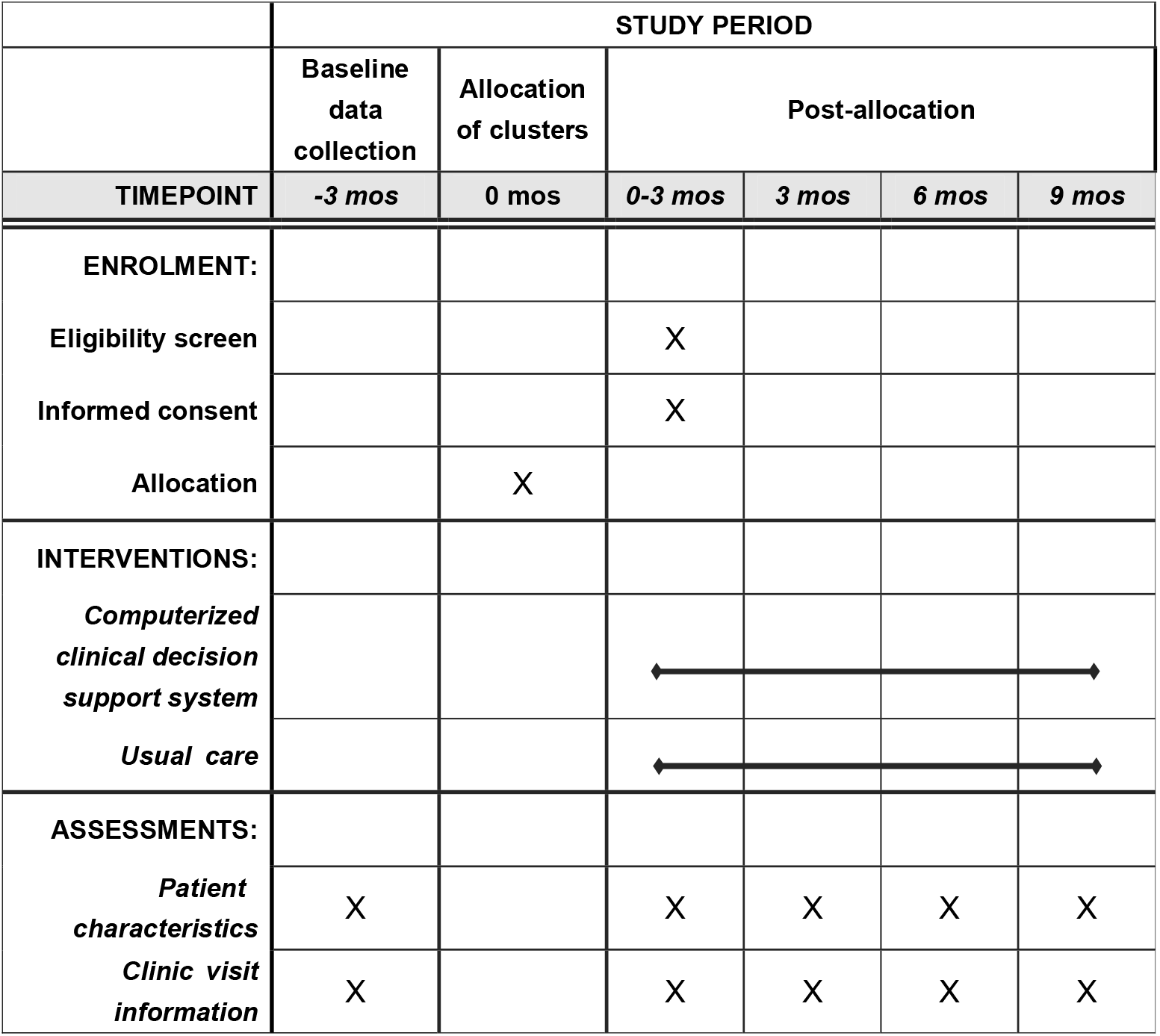
Schedule of enrollment, intervention allocation and assessment of the LIGHT trial using Standard Protocol Items: Recommendations for Interventional Trials (SPIRIT)

#### Treatment

The assignment of treatment is presented as the CDSS recommendation (class and dose). The specific agent within each class is at the physician’s discretion based on the available medications of the practice. For each case, the titration of antihypertensive medication is performed automatically by the CDSS according to the assigned treatment protocol.

#### Outcomes

The primary outcome is the difference in the change in blood pressure from baseline to 9 months between different regimens of initial therapy. Secondary outcomes include the proportion of individuals with blood pressure controlled at 9 months; the proportion of individuals with SBP <160 mm Hg and DBP <100 mm Hg at 9 months; the proportion of individuals who received monotherapy (only in Population 1), dual therapy, triple therapy, and referral at 9 months; the proportion of individuals reported to have antihypertensive drug-related side-effects; and the proportion of individuals transferred to usual care for any reasons. The exploratory outcome is the change in blood pressure from baseline to 9 months of six protocols embedded in the CDSS (A-AC-ACD; A-AD-ADC; C-CA-CAD; C-CD-CDA; D-DC-DCA; D-DA-DAC). (**Table 3**)

#### Sample size

We assume approximately 25% of the LIGHT intervention patients are in Population 1 and 75% in Population 2, with an 80% follow-up rate for the primary outcome. For each population, we estimate the detectable difference in SBP between treatment groups across a similar range of the intervention participants and statistical power. We assume that the standard deviation in SBP is σ=10 mmHg, and that the within-patient SBP correlation is R^2^ =0.2 with a maximum type II error that is Sidak-corrected for three comparisons, α=0.017.^26^ With the current number of LIGHT sites, we estimate about 2100 eligible LIGHT-ACD participants overall with complete follow-up. Under these assumptions, we estimate that for the comparisons of initial monotherapies of A, C, and D in Population 1, we will have 80% power to detect a difference of 3.5 mm Hg in SBP, and for comparisons of dual therapies of AC, AD, and CD in Population 2, we have 80% power to detect a difference of 2 mm Hg in SBP.

#### Statistical analysis

All the intervention evaluations will be performed on an intention-to-treat basis. Multiple imputation by chained equations will be used to account for missing values. For primary outcomes, we will use a linear regression model with indicators for treatment regimen and practice, and will adjust for baseline blood pressure values and imbalanced patient characteristics. Multiple Student t tests or Mann-Whitney U tests will be used; P values will be adjusted for multiple comparisons by using the Sidak method. For secondary outcomes, log-binomial regression will be used to compare groups and calculate relative risk of outcomes at 9 months. Additionally, we will perform pre-specified subgroup analyses of outcomes by age, sex, education, smoking status, and tertile of baseline blood pressure. Considering potential crossovers among treatments, we will also assess the heterogeneity of the treatment effect among per-protocol populations.

## DISCUSSION

The LIGHT trial is, to the best of our knowledge, the largest pragmatic randomized trial exploring the feasibility and effectiveness of a decision support tool to deliver high-quality care for hypertension in primary care. Moreover, by adopting a streamlined study design, we have embedded a patient-level randomized trial (LIGHT-ACD trial) into this cluster-randomized trial using an algorithm-based CDSS tool.

Our studies have several strengths. First, we have developed a usable CDSS, which can easily be integrated into the routine clinical workflow and provides tailored antihypertensive recommendations at the point of care. These features are highly correlated with the effectiveness of CDSS for improving the process of care and patient outcomes.^23, 27, 28^ As recommendations of CDSS are generated automatically by the built-in algorithm, which is developed based on current guidelines, this approach can assist primary care doctors, even those with less training, in making informed and evidence-based prescribing decisions.

Second, we have built a streamlined framework for a clinical trial that enabled us to compare the effectiveness of several guideline-based initial antihypertensive regimens. Earlier randomized clinical trials such as the ALLHAT^15^ and ACCOMPLISH^29^ trials, provided a direct comparison among several monotherapies or dual therapies. These trials are not contemporary and did not include large Asian populations, so there remain gaps in knowledge. In contrast with these standalone trials, the conduct of the LIGHT-ACD trial is embedded into the existing framework of the LIGHT trial. We incorporate a series of stepped treatment protocols into the CDSS, whereby the random allocation of recommended medication can be performed automatically by following the algorithm-consistent order at each encounter where decision support is delivered.^30^ While assessing the effectiveness of CDSS, the effectiveness of common initial antihypertensive monotherapies or dual therapies can be compared in an unobtrusive manner.^17, 29, 31, 32^

Third, the pragmatic design of both trials enables us to demonstrate real-world effectiveness of intervention/treatment with greater external validity.^33, 34^ In contrast to trials with study-specific visits, the enrollment and follow-up of patients, and the collection of outcome data in our trial are incorporated into routine clinical practice. Furthermore, the exclusion criteria are kept to a minimum to enroll a diverse spectrum of the population. These considerations improve the efficiency of trials and enhance generalizability of the study results.^34^

Fourth, the two studies are further distinguished by the efforts to build a learning decision support tool. Although the algorithm of CDSS is kept consistent during the four stages of the trial, it has the potential to be adaptively updated after the results of the LIGHT-ACD trial is available. For the ultimate implementation of the CDSS, the tool could continuously generate new knowledge in terms of the effectiveness of treatment strategies from the ongoing delivery of care, whereby the CDSS could be iteratively tested and improved by shifting the randomization ratio of stepped antihypertensive protocols toward the more effective group.^35^

Our study has some potential limitations. First, the outcomes are focused on surrogate outcomes and not clinical outcomes such as cardiovascular events. To examine the effectiveness on clinical events, a much longer trial would be required. However, it is expected that improvements in blood pressure control over time would favorably affect clinical outcomes. Second, given the nature of the CDSS, which delivers its recommendation directly to doctors, blinding was not feasible in both studies. We minimized the potential bias by using objective measures as primary and secondary outcomes. Third, due to the limited timeframe of the study, an extended follow-up was not included following the 12-month intervention to measure persistence of effects after the intervention ceases.

In conclusion, both trials should be able to provide useful evidence regarding the effectiveness of this decision support tool on improving adherence to guidelines for hypertension management in primary care, and on the comparative effectiveness of different initial antihypertensive regimens for blood pressure reduction in the real-world setting.

## Data Availability

No data is available. It is our future plan to share
the data of this study. However, we are unable to do so at this time.

## TRIAL STATUS

Protocol version 4.4 commenced on December 28, 2020. As of May 2021, 94 sites (including 7,656 patients in LIGHT and 494 participants in the LIGHT-ACD trial) have been randomized in 4 stages. Site randomization of Stage 1 was on 21 August 2019, Stage 2 on 4 December 2019, Stage 3 on 1 March 2021 and Stage 4 on 14 April 2021. All sites will complete follow-up by the beginning of 2022. In the first half of 2020, the implementation of both trials was affected by the outbreak of COVID-19. We extended the study period of the first and second stages.

## LIST OF ABBREVIATIONS

CDSS: computerized clinical decision support systems;
EHR: electronic health record;
ACE: angiotensin-converting enzyme;
ARB: angiotensin receptor blockers;
CCB: calcium channel blockers;
A: angiotensin-converting enzyme inhibitor or angiotensin receptor blocker;
C: calcium channel blocker;
D: diuretics.

## DECLARATIONS

### Ethics approval and consent to participate

The ethics committee of Fuwai Hospital approved both trials. All sites accepted this ethics approval or obtained local approval by internal ethics committees as appropriate. Given that guideline-based recommendations provided by CDSS was considered to have minimal risk for the patients, informed consent for implementing the CDSS in both trials is waived. A simple written informed consent for participants was acquired for the purpose of sending a text message as a brief medical record (e.g., blood pressure, prescriptions and follow-up reminders) after each visit.

### Consent for publication

Not applicable.

### Availability of data and materials

The datasets used and/or analysed during the current study will be available from the corresponding author on reasonable request.

### Authors’ contributions

HMK, JA, and XZ designed the study. XZ acted as the principal investigator to take responsibility for all respect of the study. JS, HMK, JA, and XZ conceived of this article. JS wrote the manuscript with further contributions from XZ, JA, HMK, JH, JW, JC, JL, XL, HZ, JL, BW, XW and YG. JH calculated the sample size and provided advice in randomization method of the study. YG performed daily data monitoring and completed all the statistical analysis. XZ, LJ, HZ, JS, XW, and BW developed the algorithm of the CDSS. All authors contributed to critical revisions and approved the final version of the article.

## Acknowledgements

We appreciate the steering committee member of the study: Robert Clarke from University of Oxford, Sharon-Lise T. Normand from the Harvard T.H. Chan School of Public Health, Frederick A. Masoudi from University of Colorado School of Medicine, Songtao Tang from the Community Health Center of Liaobu County, Wenjun Ma from the Fuwai Hospital, and Jian Xu from the Center for Chronic Disease Control of Shenzhen for their support and advice. We thank Lawrence J. Fine and George A. Mensah from National Institutes of Health for their contributions in study design. We appreciate the multiple contributions made by study teams at the National Clinical Research Center of Cardiovascular Diseases in the realms of study design and operation, particularly site management and coordination by Bo Gu, Yilan Ge, Fuyu Jing, Lei Bi, Huijun Jin, Teng Li, and Liyuan Sui, and IT development and maintenance by Shuyang Hua and Mengnan Zhu. We thank the local sites in the collaborative network for their support and data collection.

## SUPPLEMENT

**Supplement 1**. Timeline of the LIGHT trial

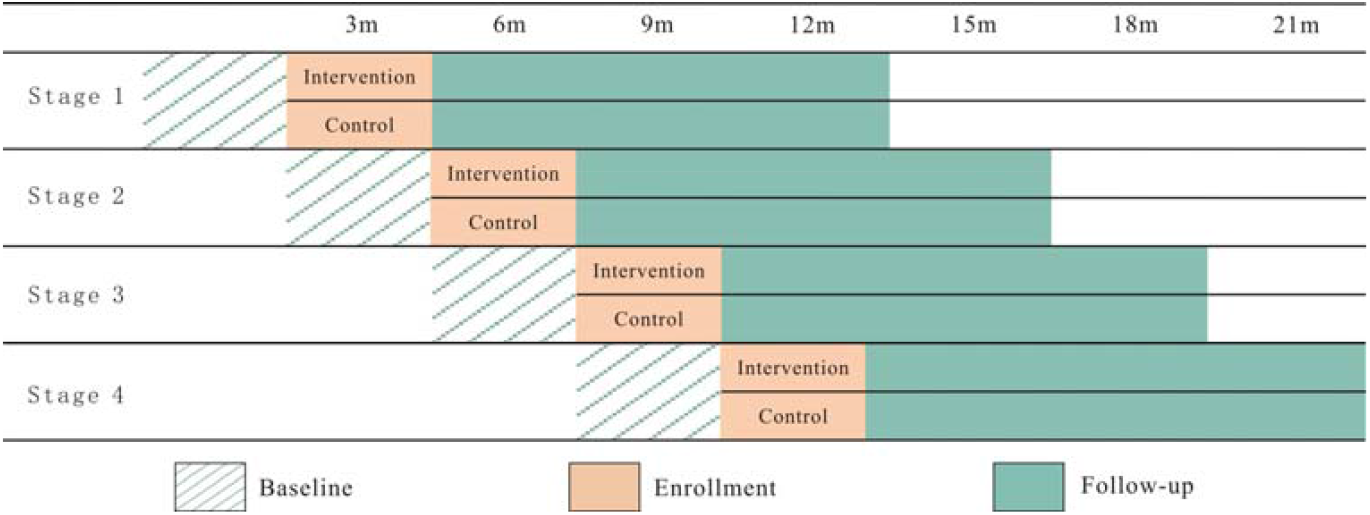

**Supplement 2**. Stratification factors for each stage in site randomization of LIGHT trial

1. Stage 1 (18 sites in Luoyang, Henan Province): baseline appropriate antihypertensive treatment rates of the site (≥median/<median) and the hospital to which the site is affiliated (Dongfang Hospital/Sixth People’s Hospital/Zhongxin Central Hosptial).
2. Stage 2 (9 sites in Zoucheng, Shandong Province): baseline appropriate antihypertensive treatment rates of the site (≥median/<median) and type of primary care practice (Fenyuan/Community health service station).
3. Stage 3 (31 sites in Shenzhen, Guangdong Province): baseline appropriate antihypertensive treatment rates of the site (≥median/<median) and geographical region (Baoan District/Nanshan District).
4. Stage 4 (36 sites in Shenzhen, Guangdong Province): baseline appropriate antihypertensive treatment rates of the site (≥median/<median) and geographical region (Luohu District/Longgang District/Futian District).

**Supplement 3**. Specification of hypertension visits

1. Scheduled or unscheduled visits for treatment of hypertension
2. Visit for diabetes, stroke, peripheral vascular disease, or other new cardiovascular diseases (i.e., chronic kidney disease, coronary heart disease, and heart failure).

**Supplement 4**. Example of CDSS recommendation

> “The recommended antihypertensive medications are C+D
>
> C (calcium channel blocker): full dose
>
> D (diuretics): half dose”
>
> Note: Full dose or half dose refer to one or half pill of commonly used antihypertensive medications respectively.

**Supplement 5**. Specifications of guideline-based antihypertensive treatment

1. Uptitrating or switching treatment for patients with inadequate blood pressure control.
  - Uptitrating or switching treatment for patients who: a) have systolic blood pressure (SBP) 140–159 mm Hg or diastolic blood pressure (DBP) 90–99 mm Hg; AND b) have established diagnosis of hypertension less than 3 months; AND c) have complications of hypertension (i.e., diabetes, chronic kidney diseases, stroke, myocardial infarction, and heart failure)
  - Uptitrating or switching treatment for patients who: a) have SBP 140–159 mm Hg or DBP 90–99 mm Hg; AND b) have established diagnosis of hypertension more than 3 months; AND c) take no antihypertensive medications.
  - Uptitrating or switching treatment for patients who: a) have SBP ≥160 mm Hg or DBP ≥100 mm Hg; AND b) take no antihypertensive medications.
  - Uptitrating or switching treatment for patients who: a) have SBP ≥140 mm Hg or DBP ≥90 mm Hg; AND b) take at least one antihypertensive medication.
2. Antihypertensive medications used in patients with specific clinical indications for their use.
  - Use of beta-blockers for patients with myocardial infarction and heart failure.
  - Use of angiotensin-converting enzyme inhibitors (ACEI) or angiotensin receptor blockers (ARB) for patients with diabetes, chronic kidney disease, myocardial infarction, and heart failure.
3. Antihypertensive medications used in patients without compelling contraindications for their use.
  - Use of diuretics for patients without gout.
  - Use of beta-blockers for patients without bradycardia (heart rate <50 beats/min).
  - Use of angiotensin-converting enzyme inhibitors (ACEI) for patients without hyperkalemia (potassium >5.5 mmol/L) or previous angioneurotic edema.
  - Use of angiotensin receptor blockers (ARB) for patients without hyperkalemia (potassium >5.5 mmol/L).
4. Antihypertensive medications used in patients without intolerance for their use
  - Use of calcium channel blockers (CCB) for patients without intolerance (e.g., CCB-induced edema) to their use
  - Use of ACEI for patients without intolerance (e.g., ACEI-induced cough) to their use
5. Use of guideline-based antihypertensive medication
  - Any antihypertensive medications beyond ACEI, ARB, beta-blocker, CCB, and diuretics not used for patients with complications.
  - Any antihypertensive medications beyond ACEI, ARB, beta-blocker, CCB, diuretics, compound reserpine triamterene, and compound reserpine) not used for patients without complications.
6. Other
  - Agents within the same class of antihypertensive medications not used at the same time.
  - Referral for patients with full dose of ACEI/ARB, CCB, and diuretics but with inadequate blood pressure control.
  - Single use of short-acting antihypertensive medications with frequency compliance to that of specified in the instruction

**Supplement 6.**
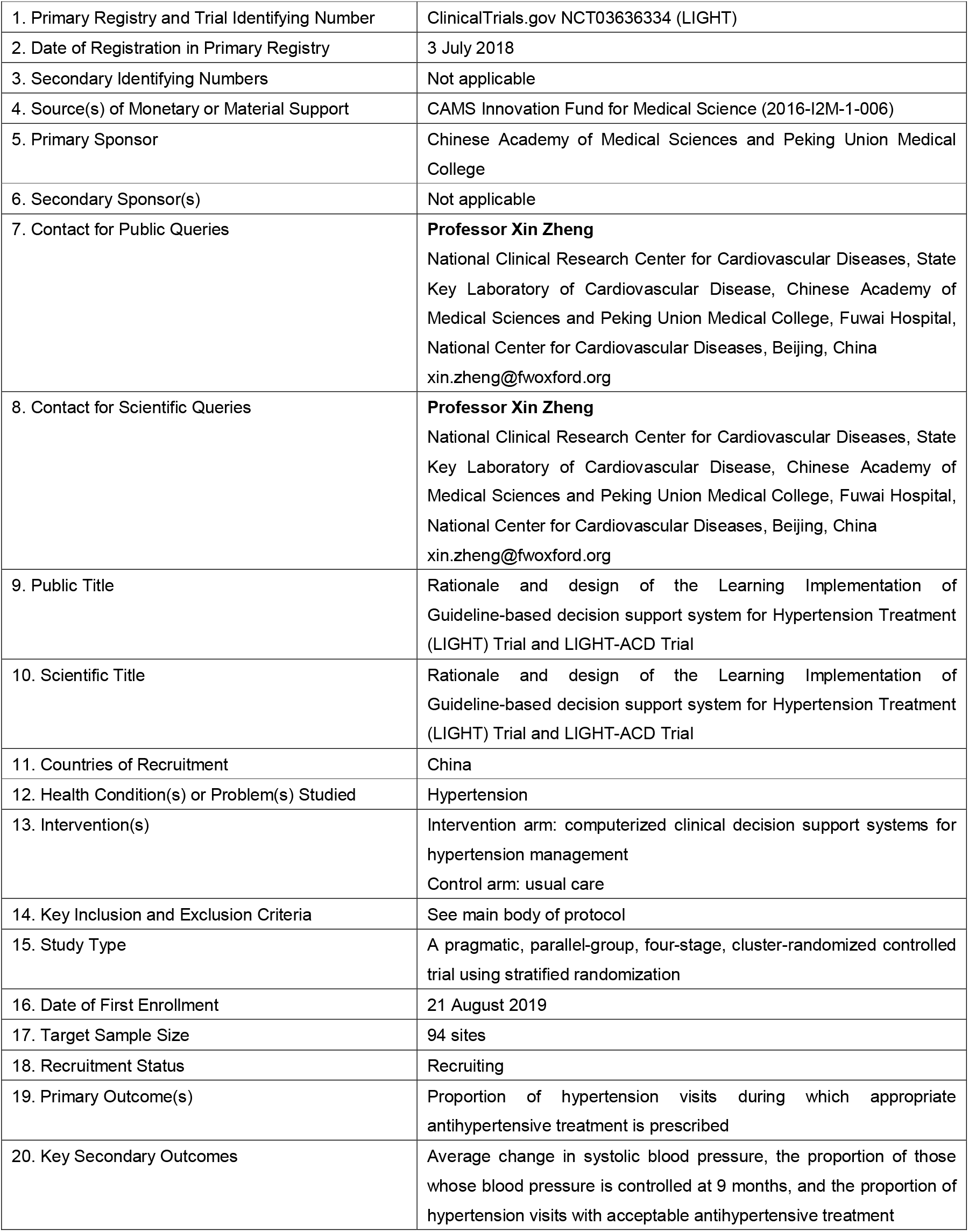
Trial registration data of the LIGHT trial

**Supplement 7.**
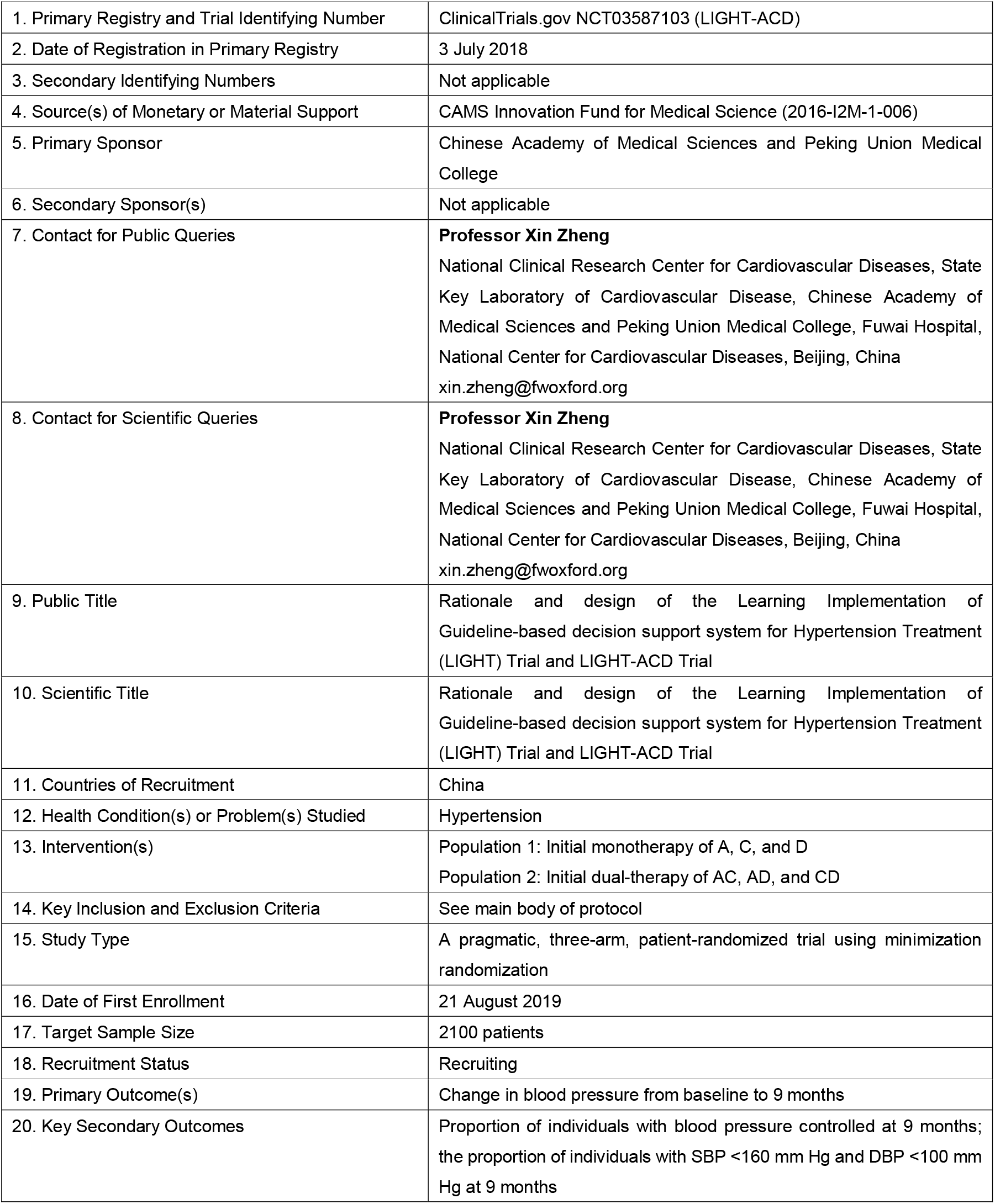
Trial registration data of the LIGHT-ACD trial

